# Evaluation of the systemic and mucosal immune response induced by COVID-19 and the BNT162b2 mRNA vaccine for SARS-CoV-2

**DOI:** 10.1101/2022.01.29.22270066

**Authors:** Olaf Nickel, Alexandra Rockstroh, Johannes Wolf, Susann Landgraf, Sven Kalbitz, Nils Kellner, Michael Borte, Jasmin Fertey, Christoph Lübbert, Sebastian Ulbert, Stephan Borte

## Abstract

**Background:** Currently used vaccines to protect from COVID-19 mostly focus on the receptor-binding domain (RBD) of the viral spike protein, and induced neutralizing antibodies have shown to be protective. However, functional relevance of vaccine-generated antibodies are poorly understood on variants-of-concern (VOCs) and mucosal immunity.

**Methods:** We compared specific antibody production against the S1 subunit and the RBD of the spike protein, the whole virion of SARS-CoV-2, and monitored neutralizing antibodies in sera and saliva of 104 BNT162b2 vaccinees and 57 individuals with natural SARS-CoV-2 infection. Furthermore, we included a small cohort of 11 individuals which received a heterologous ChAdOx1-S/BNT162b2 prime-boost vaccination.

**Results:** Vaccinated individuals showed higher S1-IgG antibodies in comparison to COVID-19 patients, followed by a significant decrease 3 months later. Neutralizing antibodies (nAbs) were poorly correlated with initial S1-IgG levels, indicating that these might largely be non-neutralizing. In contrast, RBD IgGAM was strongly correlated to nAbs, suggesting that RBD-IgGAM is a surrogate marker to estimate nAb concentrations after vaccination. The protective effect of vaccine- and infection-induced nAbs was found reduced towards B.1.617.2 and B.1.351 VOCs. NAb titers are significantly higher after third vaccination compared to second vaccination. In contrast to COVID-19 patients, no relevant levels of RBD specific antibodies were detected in saliva samples from vaccinees.

**Conclusions:** Our data demonstrate that BNT162b2 vaccinated individuals generate relevant neutralizing antibodies, which begin to decrease within three months after immunization and show lower neutralizing potential to VOCs as compared to the original Wuhan virus strain. A third booster vaccination provides a stronger nAb antibody response than the second vaccination. The systemic vaccine does not seem to elicit readily detectable mucosal immunity.

## INTRODUCTION

Starting from the pandemic spread of the coronavirus disease in December 2019 (COVID-19), global research efforts were made to identify effective vaccine candidates. Vaccines based on vectors, inactivated viruses and mRNA were licensed, whereas the latter comprises a novel immunization technology. However, the kinetics of SARS-CoV-2 antibody production and the persistence of humoral immunity following such a vaccination over time is of great interest for national health services and the management of the pandemic. Recent studies suggested that SARS-CoV-2 specific antibody production following vaccination with the BNT162b2 (BioNTech/Pfizer) mRNA vaccine is comparable to seroconversion following recovery from COVID-19.[1] During SARS-CoV-2 infection, the median time to detect circulating antibodies has been reported at 11 days after the onset of symptoms and this period will be affected by initial disease severity.[2] In patients with milder symptoms, some antibodies wane rapidly, especially IgG against the nucleocapsid protein, whereas our group and others have shown that antibodies to the spike protein and neutralizing antibodies remain detectable much longer.[3-5]

We performed a prospective study after the second BNT162b2 vaccination and assessed the longevity of vaccine-induced antibodies for three and eight months in follow-up visits and compared it with the humoral immune response after a third booster vaccination. Furthermore, we analyzed effectiveness and kinetic of BNT162b2-induced nAbs against the variants of concern B.1.351 (Beta) and B.1.617.2 (Delta) and. [6, 7] and the impact of the homologous BNT162b2 booster vaccination. Moreover, we also dissected the SARS-CoV-2 specific antibody production in individuals that were vaccinated with a heterologous ChAdOx1-S vector vaccine (AstraZeneca) and BNT162b2 mRNA vaccine (BioNTech/Pfizer) prime-boost regime. In addition to systemic immunity, mucosal immune responses are considered to be critically important in reducing SARS-CoV-2 spread. By mediation of the mucosa-associated lymphoid tissue (MALT), a strong suppression of SARS-CoV-2 transmission can be achieved due to the blockage of viral entry in mucosal cells of the oral cavity and pharynx. In saliva of recovered COVID-19 patients, IgA antibodies are detectable in high concentrations.[8] Based on the route of application, it remains unclear if homologous mRNA or heterologous mRNA-vector vaccination might elicit a partial mucosal immune response or does not provide any benefit in generating a sterile mucosal immunity.

## PATIENTS AND METHODS

### Study design and human samples

The study included two cohorts of vaccinated individuals that were followed prospectively and previously bio-banked samples from COVID-19 patients (Fig 1, Table 1). Study participants receiving homologous vaccination with the BNT162b2 mRNA vaccine (BNT/BNT, n=104, EMA fact sheet[9]) were recruited among healthcare employees at the Hospital St. Georg in Leipzig, Germany. Serum and saliva samples were collected on the day of first dose of vaccination (V1), on the day of the second dose vaccination (V2, median 21 days [IQR 21-22] after V1), and 14 to 28 days after the second dose vaccination (P1, median 42 days [IQR 42-43] after V1). We collected additional serum samples and dried blood spot samples (DBS) at time points P2 (25 individuals, 76 to 102 days after V2, median 102 days [IQR 99-109] after V1), P3 (41 individuals, [IQR 251-260] days after V2) and after booster vaccination (20 individuals, [IQR 14-16] days after P3).

**Table 1:** Demographic characteristics of vaccinees and COVID-19 cases

|  | Homologous<br>vaccinees<br>(BNT) | Heterologous<br>vaccinees<br>(AZ/ BNT) | COVID-19<br>patients |
| --- | --- | --- | --- |
| N | 104 | 11 | 57 |
| Females (%) | 68.3* | 63.6* | 43.9 |
| Median age | 41 | 31* | 51 |
| (min-max) | (20-66) | (26-37) | (32-79) |
| Median days after first dose<br>(min-max) |  |  |  |
| V2 | 21 (21-27) | 21 (20-22) |  |
| P1 | 42 (35-49) | 91 (90-92) | - |
| P2<sup>1</sup> | 102 (98-127) | - |  |
| P3 | 276 (252-286) | - |  |
| Boost | 309 (271-335) | - |  |
| Median days after the onset of<br>symptoms (min-max) | - | - | 35<br>(7-43) |
| Vaccination reaction (%): |  |  |  |
| None | 11.5 | 0 | - |
| Local<sup>2</sup> | 14.4 | 0 | - |
| Systemic<sup>3</sup> | 74.0 | 100 | - |
| severity of disease<sup>4</sup> (%) |  |  |  |
| mild (scale 2-3) | - | - | 57.9 |
| moderate/severe (scale 4-5) | - | - | 42.1 |

<sup>1</sup> blood sampling was done for 25 of 104 volunteers <sup>2</sup>pain at the vaccination site. <sup>3</sup>includes fever, chills, headache, fatigue, nausea, diarrhea, body aches, and nerve pain. <sup>4</sup>Clinical severity of COVID-19 patients was classified according to the WHO clinical progression: (2) ambulatory without limitation of activity, (3) hospitalized without oxygen, (4) hospitalized on oxygen therapy by mask or nasal prongs, (5) hospitalized receiving non-invasive ventilation or high-flow oxygen therapy. Mann-Whitney-U test was used to analyze differences in age and Fisher's exact test was applied to evaluate difference in gender (\* p<0.05, \*\*p<0.01). d = days, IQR = inter quartile range, n.s.= not significant. AZ = AstraZeneca (ChAdOx1-S), BNT = BioNTech/Pfizer (BNT162b2), V2 = three weeks after first vaccination, P1 = two weeks after second vaccination, P2 = three month after second vaccination

**Fig 1.**
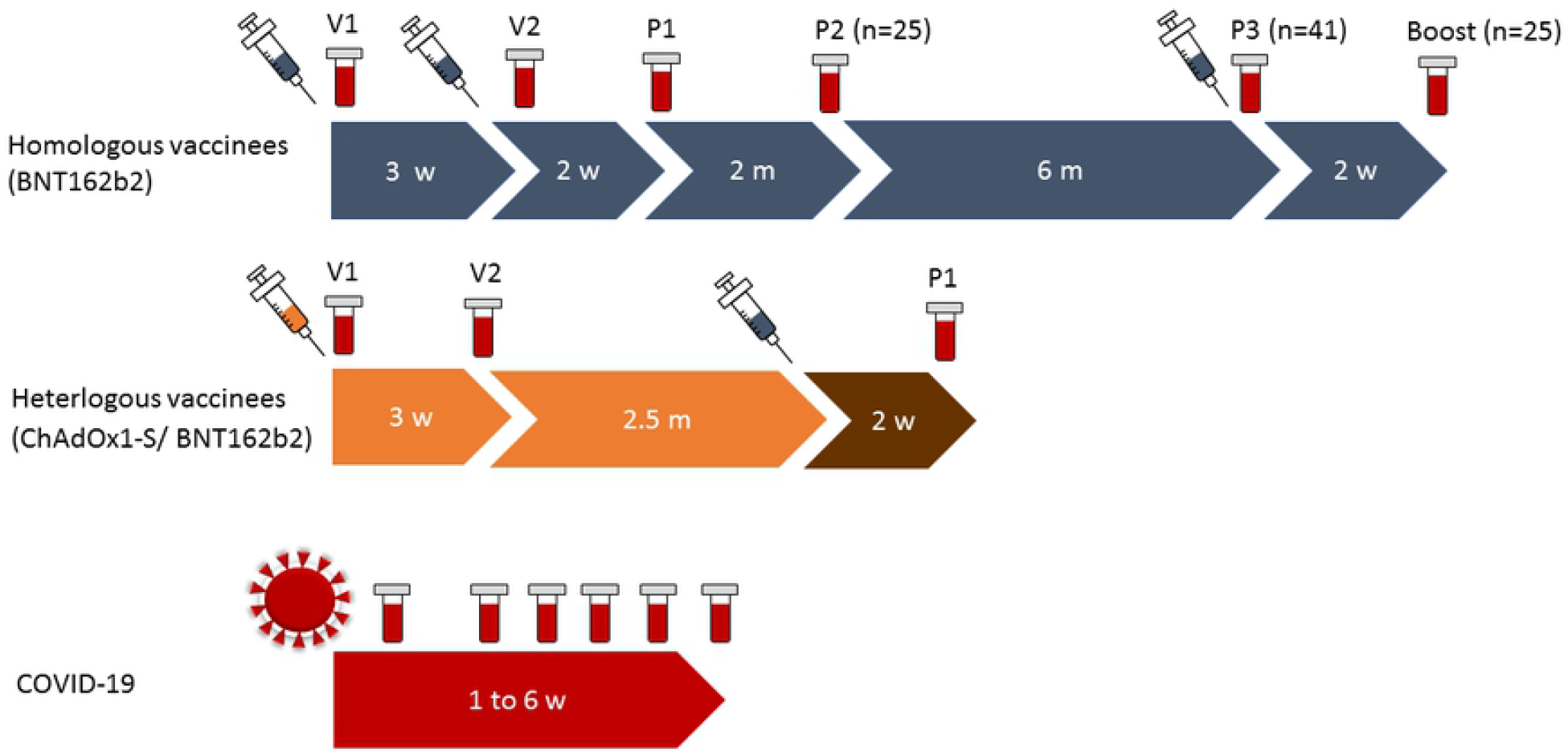
Time points of blood sampling within the different cohorts in median days (min - max) Vaccinees: V1 prior first vaccination, V2 21 days (21-27), P1 42 (35-49), P2 102 days (98-127) and P3 276 days (252-286) after first vaccination. Boost samples were taken 309 (271-335) after first vaccination. Homologous BNT162b2 vaccinees received their second vaccination 3 weeks after the first dose, whereas heterologous vaccinees received their second dose (BNT16b2) 10 weeks after the first vaccination with ChAdOx1-S COVID-19 Patients: 35 days post symptom onset (PSO) (7-43)

Another 11 study participants (63.6 % females, median age 31 [IQR 26-37]) received a heterologous ChAdOx1-S vector based prime (EMA fact sheet [10]) and BNT162b2 boost vaccination (AZ/BNT). In accordance to the homologous BNT/BNT group, serum samples were collected on the day of the first dose of ChAdOx1-S vaccination (V1), 21 days after V1 (V2, median 21 days [IQR 20-22] after V1), and 12 to 15 days after second dose vaccination (P1, median 91 days [IQR 90-92] after V1).

Inclusion into the study was independent of a previous SARS-CoV-2 infection. The ethics committee of the Saxonian medical chamber approved the study (registry number EK-allg-37/10–1) and informed consent was obtained from all volunteers.

As reference, 57 PCR-confirmed COVID-19 patients were included, which were treated between March 3 and November 11, 2020, at the Department of Infectious Diseases/Tropical Medicine, Nephrology and Rheumatology of the Hospital St. Georg. Serum samples were collected between 7 and 43 days (Median 35 days, IQR 20-44) after symptoms onset. COVID-19 patients were stratified into two groups according to the WHO clinical progression scale *(World Health Organization 2020 ordinal scale for clinical improvement)*: (1) “mild”, scale values 2 or 3 and (2) “moderate/severe” with scale values 4 or 5. For 29 of 34 COVID-19 patients with moderate/severe progression, saliva was collected (Table 1).

### Commercial assays for the detection of antibodies against S1 and Nucleocapsid

All serum samples were tested for IgG against SARS-CoV-2 S1 (Anti-SARS-CoV-2-QuantiVac-ELISA, S1 Quant IgG; cut-off ≥25.6 BAU/ml) and for IgA against SARS-CoV-2 S1 (S1 IgA, Euroimmun, Lübeck, Germany; cut-off ratio ≥0.8). Samples above detection limit for S1 Quant IgG were pre-diluted 1:10 and 1:50 in sample buffer. In addition, baseline sera were screened for IgG antibodies against SARS-CoV-2 nucleocapsid (Virotech, Rüsselsheim, Germany; cut-off ≥11 VE/ml).

### In-house developed enzyme-linked immunosorbent assays

Detection of SARS-CoV-2 inactivated whole virion (IWV) IgG-antibodies and SARS-CoV-2 RBD polyvalent IgGAM-antibodies was performed according to Rockstroh *et al*. 2021.[3] Briefly, Nunc PolySorp plates were coated with 1.5 µl per well of inactivated SARS-CoV-2 wt viral particles and 250 ng/well of RBD protein in 100 µl per well of carbonate coating buffer (15 mM Na2CO3, 7 mM NaHCO3 pH 9.6) overnight at 4°C. RBD protein (AA residues 329-538 of spike protein, strain Wuhan-Hu-1) was expressed in *Drosophila S2* cells and purified from cell culture supernatants with tandem immobilized metal affinity and size exclusion chromatography using the ÄKTA pure 25 l chromatography system (GeHealthcare). SARS-CoV-2 viral particles were purified from infectious cell culture supernatants by ultracentrifugation on a 30 % sucrose cushion in MSE buffer (10 mM MOPS, pH 6.8, 150 mM NaCl, 1 mM EDTA) at 25,000 rpm for 3.5 h and 4°C. The pellet was resuspended in MSE buffer and chemically inactivated with 0.1 % beta-propiolactone at 22°C. Sera (diluted 1:100) were incubated for 1.5 h at room temperature and binding antibodies were detected using a HRP-conjugated secondary goat anti human IgG antibody (Dianova, 1:20,000) or goat anti human IgG+IgM+IgA H&L antibody (Abcam, 1:10,000) for 1 h at room temperature. TMB substrate (Biozol) was added after a final wash step and incubated for 25 minutes before the reaction was stopped with 1 M H2SO4. Absorbance was detected at 450 nm with 520 nm as reference in a microplate reader (Tecan). The cut-off values were determined for each antigen individually and were validated using 100 pre-pandemic serum samples. All measurements were performed at least in duplicates.

### SARS-CoV-2 viral stocks

SARS-CoV-2 wildtype virus (wt) (isolate BetaCoV/Germany/BavPat1/2020, obtained from the European Virus Archive Global, EVAg)[11], SARS-CoV-2 B.1.1.7 (isolate MUC1-IMB1-CB, kindly provided by Klaus Überla from the Institute of Clinical and Molecular Virology, University of Erlangen-Nürnberg) and SARS-CoV-2 B.1.351 (SARS-CoV-2/human/Germany/LE-B14HXA2/2021 kindly provided by Corinna Pietsch from the Institute of Virology, University Hospital Leipzig) were propagated in VeroE6 cells. Cells were grown to a confluence of approx. 80 - 90 % and were infected at a MOI of 0.001 in Dulbecco’s modified Eagle’s medium (DMEM), supplemented with 2 % FCS and 1 % Pen/Strep. They were incubated for 48 h at 37°C with 5 % CO_2_ until cytopathic effect (CPE) was visible. Virus containing supernatants were centrifuged at 4000 g for 10 min at 4°C and then stored at -80°C until use. Viral titers were determined using a focus-forming assay. All viral stocks were sequenced to verify their spike protein sequences and expected mutation sites.

### SARS-CoV-2 neutralization assay

Focus reduction neutralization assays (FRNT) were performed according to Rockstroh *et*. al. 2021.[3] Briefly, heat-inactivated human serum samples were serially diluted in DMEM without FCS from 1:2.5 to 1:5120 and incubated with 50-150 focus forming units of SARS-CoV-2 wt, B.1.1.7 or B.1.351 for 1 h at 37°C before addition to confluent Vero E6 monolayers in 96-well plates. After an incubation of 1 h at 37°C, supernatant was removed, cells were washed with PBS, overlaid with 1 % Methyl cellulose in DMEM with 2 % FCS and incubated for 24-26 h at 37°C in 5 % CO2. Cells were fixed with 4% paraformaldehyde in PBS, permeabilized and blocked with Perm-Wash buffer (0.1% saponin, 0.1% BSA in PBS). SARS-CoV-2 focus forming units were stained using a monoclonal rabbit anti-S1 antibody (CR3022, abcam, 1:1,000) and a secondary goat anti-rabbit IgG HRP-conjugated antibody (Dianova, 1:1,000). After the addition of TrueBlue substrate (Seracare), spots were counted with an ELISpot reader (AID Diagnostika). FRNT_90_ titers were determined as the reciprocal of the last dilution providing a minimum of 90 % neutralization of focus forming units in comparison to the virus control without serum. A positivity cut-off of FRNT_90_ ≥5 was determined with negative reference sera, data not shown.

### Collection and detection of SARS-CoV-2 specific IgA in saliva

Saliva was collected using the Oracol Saliva Collection System (Oracol, Worcester, U.K.) according to the manufacturer’s specification. Aliquots were stored at -80°C in Protein Low-Bind microtubes (Eppendorf, Hamburg, Germany) until further use. Samples were centrifuged at 10,000 g for 5 min and the supernatant was collected. 25 µl of saliva was mixed with equal amounts of LEGENDplex assay buffer and S1- or RBD-coated beads in a 5 ml polypropylene FACS tube, sealed and incubated overnight at 7°C in the dark. Subsequently, the bead mixture was washed with 1 ml of LEGENDplex wash buffer at 250 g for 5 min and incubated with 25µl of Streptavidin-conjugated anti-IgA detection antibody for 60 min on an orbital shaker at 800 rpm, followed by the addition of 25 µl of Streptavidin-PE conjugate for another 30 min (all from BioLegend, SanDiego, CA, U.S.). After another wash step, beads were resuspended in 500 µl of BD sheath fluid and analyzed using a BD FACS Lyric flow cytometer (BD Biosciences, San Jose, CA, U.S.) with PMT voltage settings adapted to discriminate beads specific for Spike S1- and Spike RBD-specific IgA antibodies. Binding of IgA antibodies was evaluated as median fluorescence signals on detected beads.

### Study design and human samples

For comparison of surrogate neutralizing antibody concentrations to SARS-CoV-2 in dried blood spot samples DBS, we included samples of 63 COVID-19 patients (53.9% female, median age 61 years, IQR 48-72) collected between 60 and 319 days (Median 177 days, IQR 146-258) after disease onset.

### Validation of a dried blood spot based surrogate assay for SARS-CoV-2 nAbs

Given the superior value and importance of a correlate of serological neutralizing activity in SARS-CoV-2 vaccinated individuals and COVID-19 patients, as well as to monitor seasonal booster vaccinations, we intended to develop and validate a simple diagnostic platform using DBS. This ACE-2 competitive-binding assay uses non-purified serum eluates from DBS and is scalable in a 96-well format for automated analysis on a suitable reader system capable of detecting fluorophore-conjugated microbeads.

With this approach, we could verify neutralizing surrogates both in mild and severe COVID-19 courses, as well as in BNT162b2-vaccinated individuals, with the latter two showing comparable levels as previously shown in the assessment of neutralizing antibodies using our focus reduction neutralization assay (FRNT) with serum samples. Therefore, we compared FRNT-suggested nAb titers in matched serum samples with calculated concentrations of neutralizing surrogate antibodies in DBS and found a high degree of correlation (S3).

### Detection of surrogate neutralizing antibodies to SARS-CoV-2 in saliva and dried blood spot samples

Dried blood spot samples (DBS) were generated by applying peripheral venous blood on a GE Healthcare / Whatman 903 filter paper (Little Chalfont, U.K.) allowed to dry at room temperature for at least 4 hours. DBS were stored at 7°C until further use. 2 × 3.2 mm dots were generated using a PerkinElmer Wallac punching device and were sorted into 96-well 0.45 µM PVDF plates (Merck Millipore, Billerica, MA, U.S.) with plate outlets sealed with a foil. 70 µl of LEGENDplex assay buffer was applied and plates were sealed on top, centrifuged at 1000 g for 2 min and stored at 7°C overnight. The outlet foil then was removed and the DBS suspension was drained from the plates by centrifugation at 1000 g for 5 min into a 96-well Protein LowBind plate (Eppendorf, Hamburg, Germany). Saliva samples were collected, stored and prepared as depicted above. 50 µl of DBS suspension or saliva was combined in a 5 ml polypropylene FACS tube together with 25 µl of SARS-CoV-2 neutralizing antibodies and 25 µl of SARS-CoV-2 neutralizing assay beads B3 (all from BioLegend, SanDiego, CA, U.S.) and kept on an orbital shaker for 120 min at 800 rpm. Subsequently, the bead mixture was incubated with 25 µl of Streptavidin-PE conjugate for another 30 min and finally washed with 1 ml of LEGENDplex wash buffer at 250 g for 5 min. Beads were resuspended in 500 µl of BD sheath fluid and analyzed using a BD FACS Lyric flow cytometer with PMT voltage settings adapted to detect beads specific for the ACE2 neutralization target. Binding of neutralizing antibodies inversely correlates with the median fluorescence signal detected on beads.

### Statistical analysis

SPSS version 21 (IBM, Armonk, NY, USA) and GraphPad PRISM version 6 (GraphPad Software, San Diego, CA, USA) were used for statistical calculations and generation of figures. Statistical tests were calculated as paired or unpaired one-way ANOVA with Holm-Sidak’s multiple comparison test for the analysis of follow-up samples or vaccine and infection induced antibodies, respectively. Fisher’s exact test was applied for comparison of categorical variables.

## RESULTS

### Serological characterization of BNT162b2 vaccinees and COVID-19 patients

Among 104 individuals receiving a homologous BNT162b2 vaccination we analyzed S1, RBD, inactivated whole virion (IWV) and neutralizing antibodies and drew comparisons to 57 COVID-19 patients with mild and severe courses serving as reference group (Table 1,Fig 2 A-E).[3] SARS-CoV-2 neutralizing as well as S1, RBD and IWV binding antibodies were detectable in most individuals three weeks after the first vaccination. Two weeks after the second dose, nAb titers and binding antibodies to all tested antigens were significantly increased. At this stage, nAb titers (Median FRNT_90_ = 320) as well as IWV IgG antibody signals were comparable to those induced by natural mild COVID-19 but did not reach the high levels of patients having severe COVID-19 courses (4- and 1.3-fold decrease for nAb and IWV IgG respectively) (Fig 2 A and E). In contrast, RBD IgGAM and S1 IgA antibody levels corresponded to those induced after severe COVID-19 and were even significantly exceeded by vaccine-induced S1 IgG antibodies (3.5 fold increase). For some individuals, IgG antibodies against the inactivated whole virion of SARS-CoV-2 (IWV) were detected even before the first vaccination.

**Fig 2.**
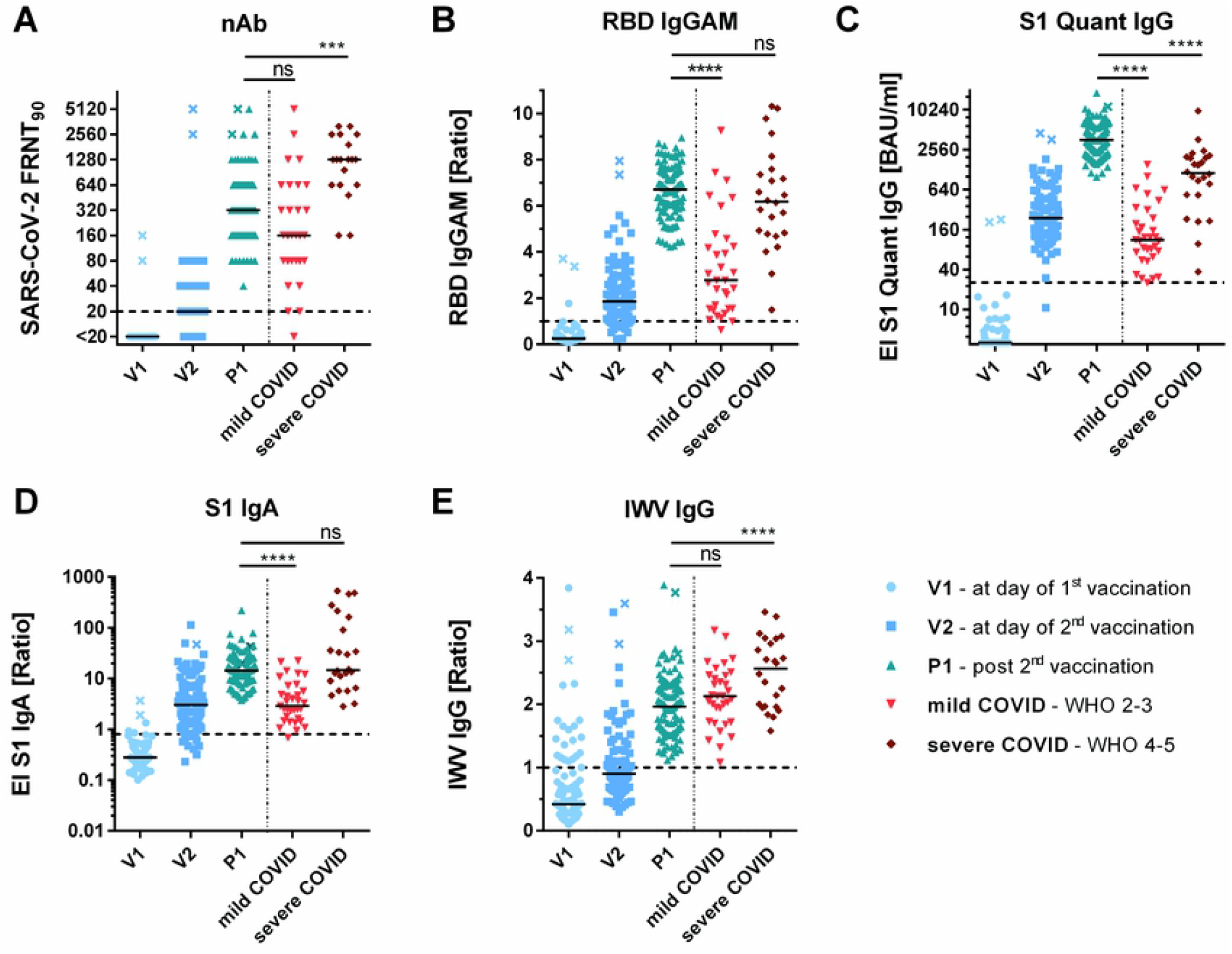
BNT162b2 vaccination induced antibodies in comparison to SARS-CoV-2 infection induced antibodies The presented data shows antibody ratios before first (V1), 3 weeks after first (V2), and 2 weeks after second vaccination (P1), as well as antibodies in sera of patients after mild and severe COVID-19 course. (**A**) Reciprocal titers of SARS-CoV-2 neutralizing antibodies (nAb) were measured in focus reduction neutralization assay with 90 % inhibition (FRNT_90_). (**B**) RBD IgGAM signals determined as sample/cut-off ratios (**C**) S1 quant IgG antibodies were quantitatively measured in binding antibody units per milliliter (BAU/ml). (**D-E**) S1-IgA and inactivated SARS-CoV-2 whole virus IgG signals (IWV) were determined as sample/calibrator ratios The horizontal dotted lines represent positivity cut-offs. Statistical analysis was performed with ordinary 1-way ANOVA, Holm-Sidak’s multiple comparison test * = *p*<0.05, ** = *p*<0.01, *** = *p*<0.001, **** = *p*<0.0001, ns = not significant x-marked data points represent vaccinees with a previous SARS-CoV-2 infection

The correlation between BNT162b2-induced S1 IgG, RBD IgGAM antibody signals and nAb titers using Spearman’s rank coefficient is presented in S1 Fig. Herein, S1 IgG and RBD IgGAM antibodies indicated the strongest correlation to SARS-CoV-2 nAbs (*r*=0.93) (Fig 3 A-B), whereas IWV and S1-IgA correlation coefficients ranged between *r*=0.722 and 0.817 (S1 Fig C-D).

**Fig 3.**
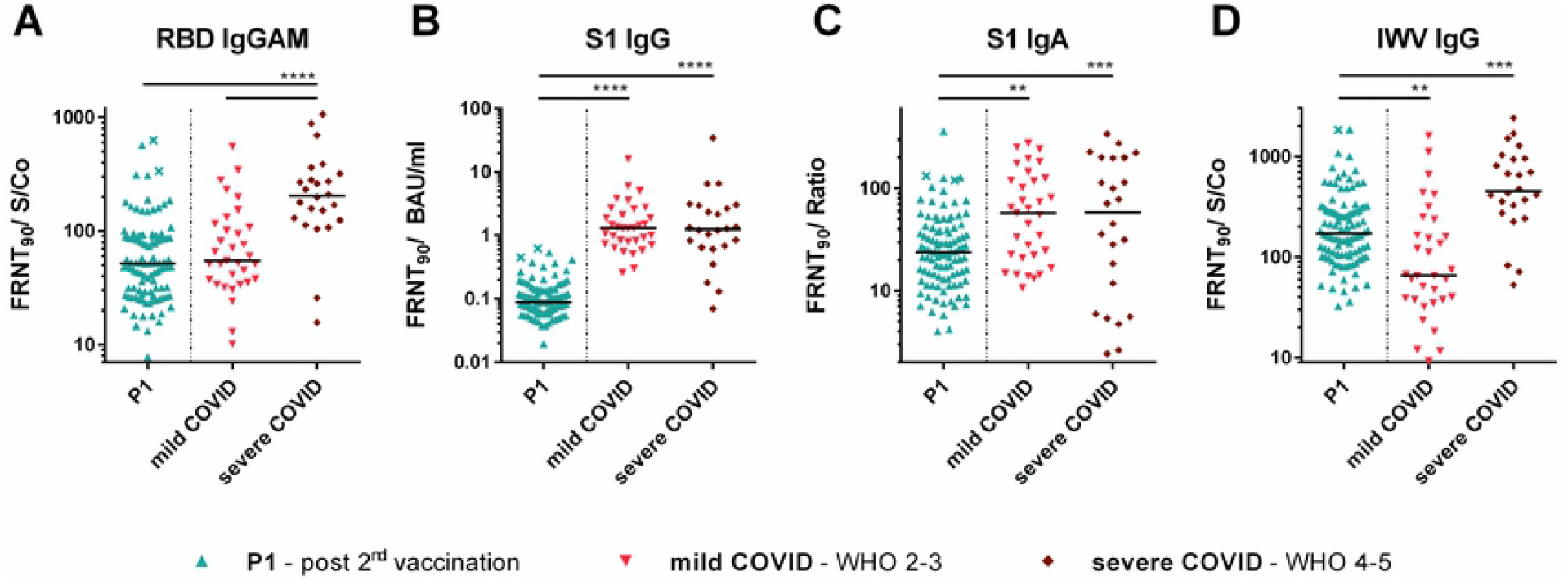
Ratios between SARS-CoV-2 neutralizing antibody titers and (**A, C, D**) RBD IgGAM, S1 IgA, IWV in sample/calibrator ratios and (**B**) S1 quant IgG in BAU/ml. Sera of vaccinees two weeks after second vaccination (P1) were compared with sera of COVID-19 patients with mild and severe courses according to WHO score (2-3 mild and 4-5 severe). Statistical analysis was performed with an ordinary 1-way ANOVA, Holm-Sidak’s multiple comparison test * = *p*<0.05, ** = *p*<0.01, *** = *p*<0.001, **** = *p*<0.0001, ns = not significant.

The heterologous AZ/BNT vaccination cohort showed similar nAb titers after the first dose in comparison to BNT162b2 vaccinees (S2 Fig). However, after the second vaccination significantly higher nAb titers (4 fold increase) were detected compared to the homologous mRNA vaccine group.

### Comparison of binding and neutralizing SARS-CoV-2 antibodies

Ratios of binding to neutralizing antibodies were calculated to compare the proportion of neutralizing antibodies induced by vaccination to convalescent individuals in each test (Fig 3). Vaccinees presented a significantly lower proportion of neutralizing to S1 binding antibodies in comparison to the COVID-19 group (10-fold and 5-fold lower for S1 IgG and S1 IgA respectively). Similarly, this ratio was lower for RBD binding IgGAM antibodies compared to patients after severe COVID-19 but was found comparable to the mild COVID-19 group. For IWV-IgG the neutralizing proportion of binding antibodies was higher than in patients with mild COVID-19 courses but below the median of severe courses.

### Long term kinetics of antibody titers

The kinetics of antibody abundance varied greatly, with a mean reduction of nAb by 3.3 fold at P2 and 9.7 fold at timepoint P3 compared to timepoint P1 two weeks after second vaccination. (Fig 4 A) RBD IgGAM ratios decreased 1.8 fold after three month and 4.0 fold after eight month (Fig 4 C). S1 antibody concentrations fall 5.7 fold at timepoint P2 and 36.6 fold at P3 (Fig 4 C). Smallest differences were observed in the IWV IgG antibody signals with a 1.1 fold mean reduction after three months and a 2.2 fold mean reduction after eight month (Fig 4 D).

**Fig 4.**
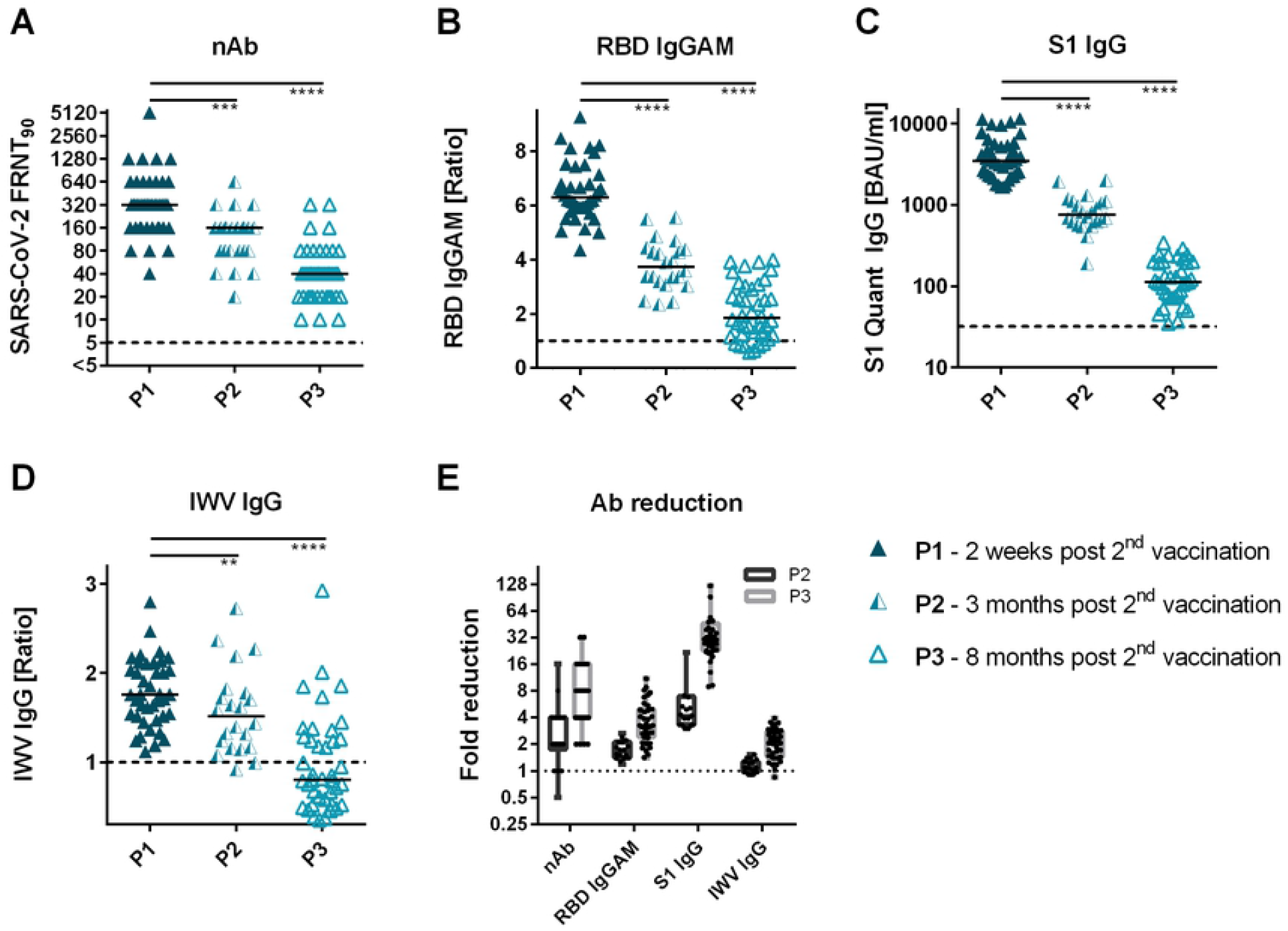
BNT162b2-induced antibody levels 2 weeks, 3 and 8 month after second vaccination. (A) Reciprocal titers of SARS-CoV-2 neutralizing antibodies were measured in focus reduction neutralization assay with inhibitor titer 90% (FRNT90). (B) RBD IgGAM signals were determined as sample/calibrator ratios (C) S1 IgG antibodies were quantitatively measured in binding antibody units per milliliter (BAU/ml). (D-E) S1 IgA and IWV (SARS-CoV-2 inactivated whole-virion) titers were determined as sample/calibrator ratios. (F) Fold reduction of SARS-CoV-2 neutralizing antibody titers, RBD IgGAM, S1 IgG, S1 IgA and IWV IgG antibody signals in follow-up vaccine sera calculated as ratio of value on timepoint P1 to timepoint P3.

### Neutralizing effect of BNT162b2-induced antibodies on B.1.617.2 and B.1.351 VOCs

Neutralizing antibody titers were decreased towards VOCs in almost all tested individuals and cohorts when compared with wildtype SARS-CoV-2 (Fig 5). A 5.1- and 11.5-fold mean titer decrease was observed with the B.1.617.2 and B.1.351 variant, respectively. With 6.2-fold and 7.7-fold a similar nAb mean reduction was observed for both variants at P2. After eight month nAb titers for B.1.617.2 showed a 6.2 fold and for B.1.351 a 4.1 fold decrease. Compared to the homologous BNT/BNT vaccination group and Covid 19 convalescent group, the decrease of nAbs against VOCs was much smaller (2.5- to 3.5-fold decrease) in the heterologous AZ/BNT vaccination cohort (S3 Fig).

**Fig 5.**
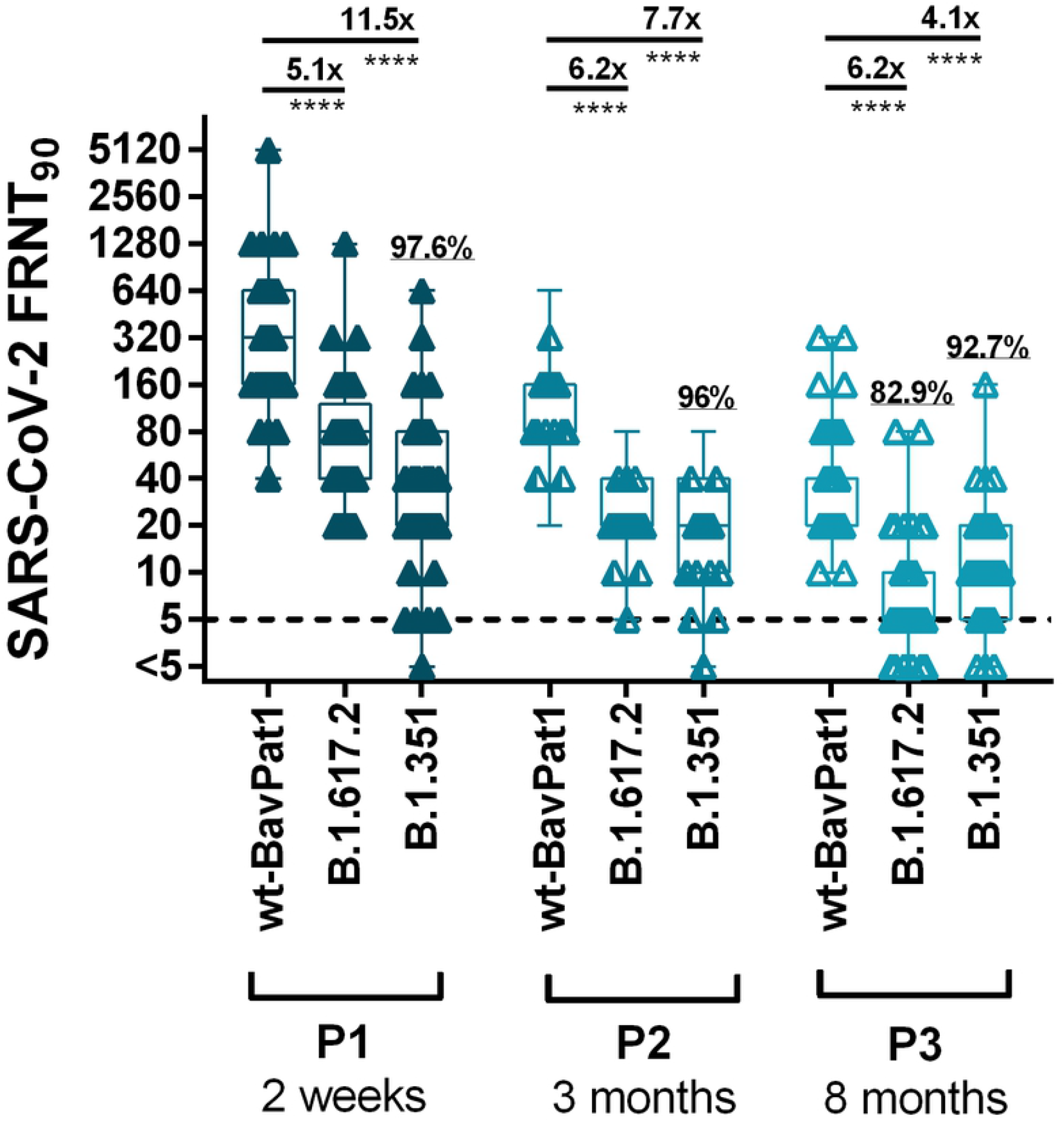
BNT162b2- and infection induced reciprocal SARS-CoV-2 nAb titers on the wild-type strain wt-BavPat1 and the VOC strains B.1.617.2 and B.1.351 at timepoint P1 to timepoint P3. The dotted line indicates the limit-of-detection at a titer of 1:5. FRNT90: focus reduction neutralization titer at 90% virus inhibition; results plotted as reciprocal values. Mean neutralizing titer reductions of SARS-CoV-2 wt to VOC-nAb are depicted above the continuous lines * = *p*<0.05, ** = *p*<0.01, *** = *p*<0.001, **** = *p*<0.0001, ns = not significant

### Impact of booster vaccination to nAb titer level on wildtype, B.1.617.2 and B.1.351 strains

Two weeks after booster vaccination nAb titers are significantly higher compared to time point P1 after second vaccination. The median increase reaches from 6.1 fold for wildtype variant to 26.2 fold in B.1.351 (Fig 6 A). The comparison between time point P3 and boost shows median increase ranges between 76.3 fold and 138 fold (Fig 6 B).

**Fig 6.**
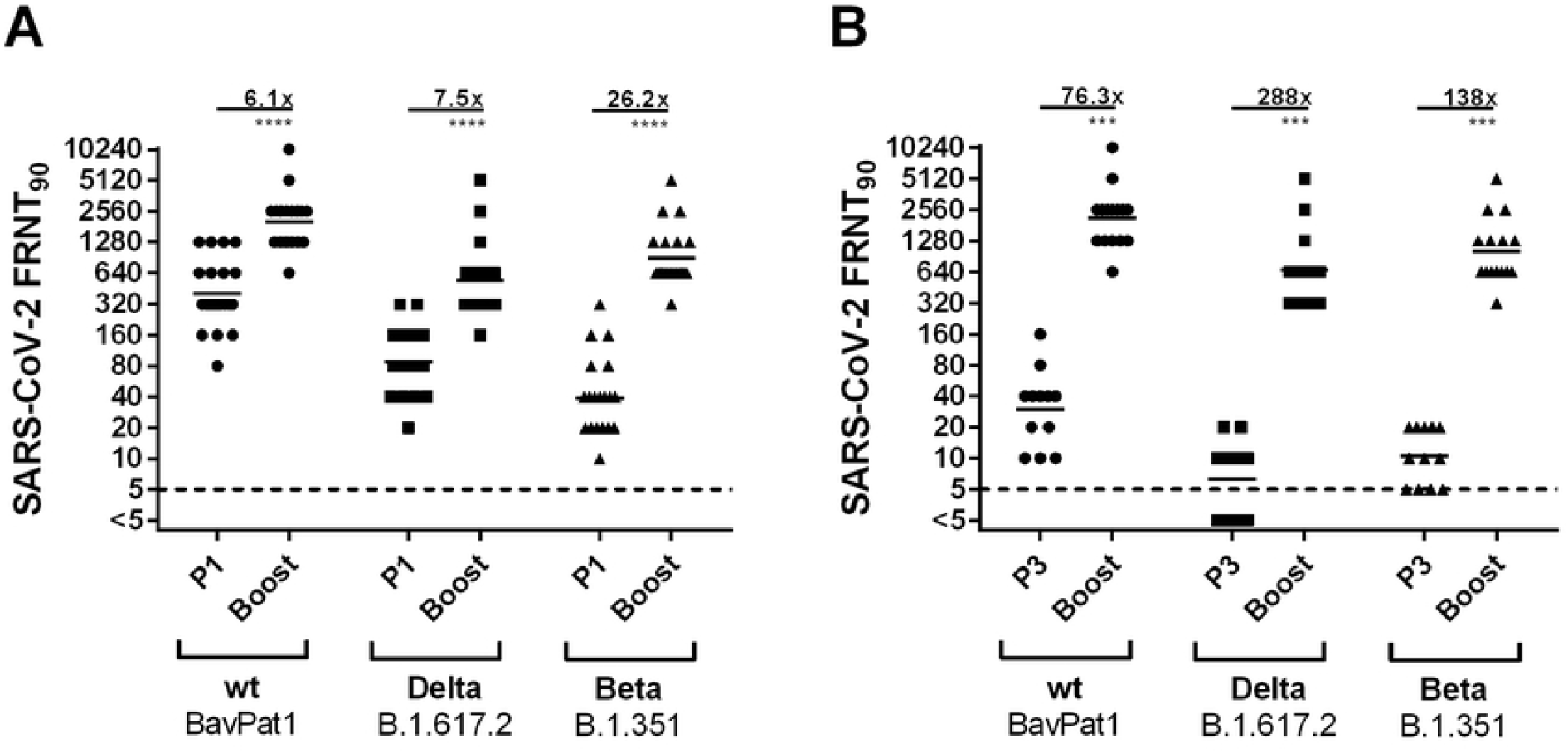
BNT162b2-induced reciprocal SARS-CoV-2 nAb titers on the wild-type strain wt-BavPat1 and the VOC strains B.1.617.2 and B.1.351. (A) Nab titers were compared two weeks after second and third vaccination. (B) Comparison of nAbs 8 month after second and two weeks after third vaccination. The dotted line indicates the limit-of-detection at a titer of 1:5. FRNT90: focus reduction neutralization titer at 90% virus inhibition; results plotted as reciprocal values. Mean neutralizing titer reductions of SARS-CoV-2 wt to VOC-nAb are depicted above the continuous lines * = *p*<0.05, ** = *p*<0.01, *** = *p*<0.001, **** = *p*<0.0001, ns = not significant

### Validation of a dried blood spot based surrogate assay for SARS-CoV-2 nAbs

FRNT-suggested nAb titers in serum samples were highly comparable (r=0.664) (S4 Fig) to concentration of neutralizing surrogate antibodies in DBS, as further outlined in the Supplementary material.

### Detection of SARS-CoV-2 targeting antibodies in saliva samples

Saliva samples from vaccinees and COVID-19 patients with moderate or severe courses were assessed for S1- and RBD-specific IgA antibodies using a sensitivity-trimmed bead-based flow-cytometric assay. Based on interquartile range calculation, no increase of IgA production at V2 or P1 was observed in the group of BNT162b2-vaccinated individuals. Similar results were obtained for the AZ/BNT group (data not shown). In contrast, most COVID-19 patients had detectable salivary IgA towards SARS-CoV-2 antigens after 15-30 days after the onset of symptoms (Fig 7).

**Fig 7.**
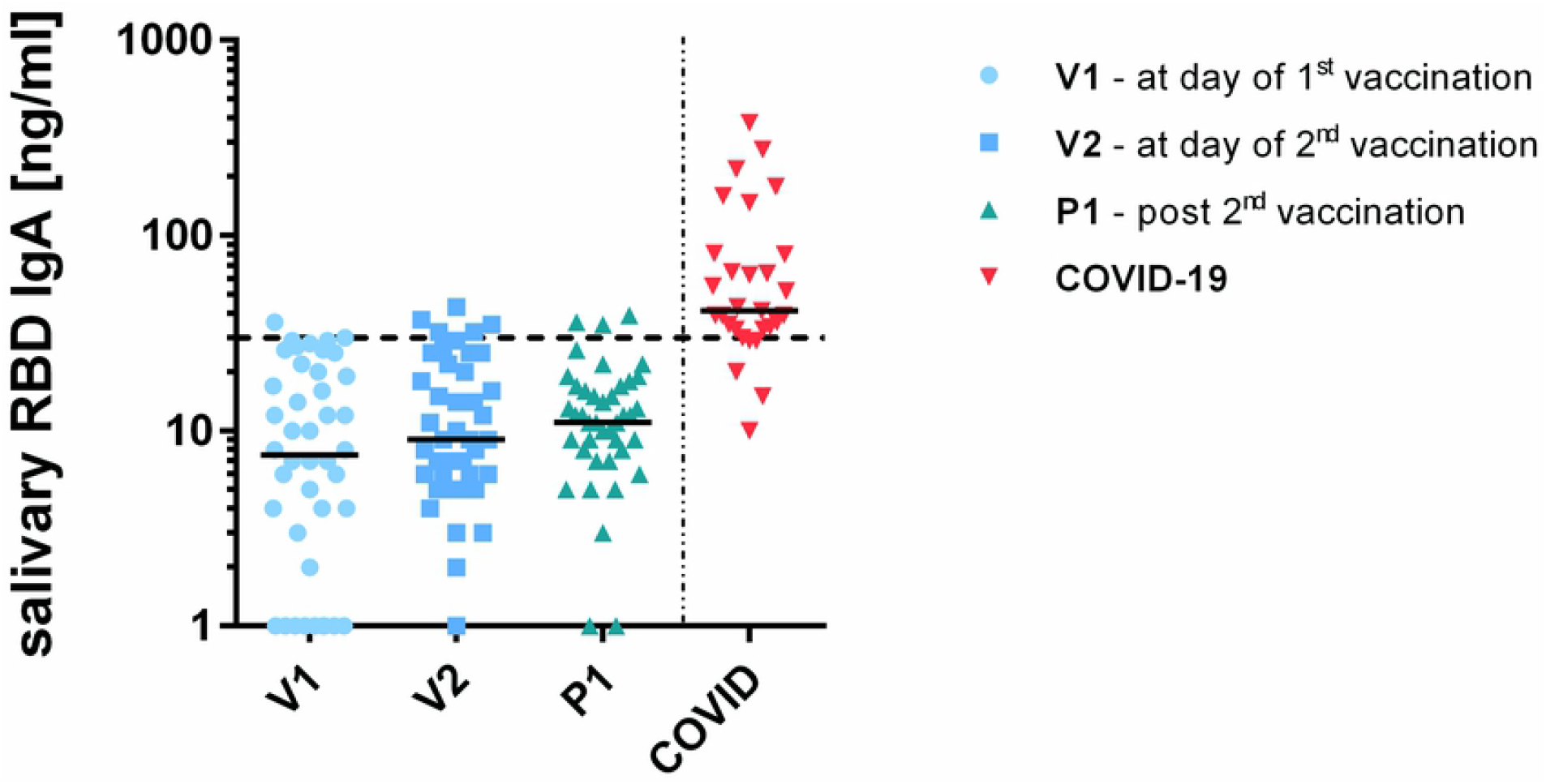
Salivary IgA specific to SARS-CoV-2 RBD in individuals prior first **(V1)**, three weeks after first **(V2)** and to weeks after second vaccination **(P1)** in comparison with COVID-19 patients. The dotted line indicates the 95% interquartile range for vaccinees.

## DISCUSSION

### Impact of vaccination against SARS-CoV-2 on antibody formation

Vaccination with mRNA- or vector-based vaccines represents a milestone in combating the SARS-CoV-2 pandemic. To better understand their protective effects, we assessed the antibody formation quantitatively and functionally after vaccination in comparison with natural SARS-CoV-2 infections. Three weeks after the first vaccination with BNT162b2, in half of the subjects nAbs could be detected, which is of note with regard to recent studies linking early levels of nAbs with protection against SARS-CoV-2.[12] This suggests a presumably protective effect already after the first vaccination with the BNT162b2 mRNA vaccine. Two weeks after the second dose, all individuals developed high S1- and RBD-binding as well as SARS-CoV-2 nAb titers. These results are in concordance with similar observations in studies on the mRNA-1273 vaccine.[13] Interestingly, an even stronger nAb production was observed in the group of heterologous ChAdOx1-S/BNT162b2 vaccinated individuals. This indicates that a combination of different SARS-CoV-2 vaccine classes leads to stronger humoral immune response which may result in a better protective effect, as has also been shown by other studies. [14-17].

### Long-term kinetics of SARS-CoV-2 antibody response after BNT162b2 vaccination

We have observed significant reduction of vaccine-induced antibody levels three months after the second vaccination. Interestingly, S1 quant IgG and IgA antibodies decreased strongly, whereas nAb levels and RBD-GAM dropped to a lesser extent.[18] Thus, the S1 quant IgG ELISA does not seem to be an optimal diagnostic choice for determining longevity of humoral immune responses after SARS-CoV-2 mRNA vaccination. Therefore, we propose the use of RBD-IgGAM determination as a rapid and simple surrogate marker to estimate the levels of nAbs after SARS-CoV-2 mRNA vaccination. One explanation for a better correlation between RBD-IgGAM and nAbs could be a better RB-Domain presentation in the vaccine antigen. Considering that a broader nAb production against the RBD region was observed in vaccinees compared to COVID-19 patients.[19] Furthermore, as an alternative with potential for the easier handling of study samples we introduced a new method to detect surrogate nAbs from dry blood spot cards, and results from this assay highly correlated with the nAb data obtained in a classical virus neutralization test. Of note, for any serological test addressing protective responses, suitable cut-off levels reflecting the biological relevance would have to be determined within future studies in larger patient cohorts.

As an observed decrease in SARS-CoV-2 antibody titers within a short diagnostic interval might lead to the demand of booster vaccination doses, it is important to also investigate the cellular and memory immune responses in order to provide a full picture of SARS-CoV-2 protection. Data from recovered COVID-19 patients implies the presence of memory B- and T-cells in almost all individuals up to eight months after SARS-CoV-2 infection.[5, 20]

### Vaccination or SARS-CoV-2 infection induced nAbs against SARS-CoV-2 VOCs

The BNT162b2 vaccine was designed using the spike gene sequence of the original Wuhan SARS-CoV-2 wildtype virus. During the pandemic spread of SARS-CoV-2, mutations created many viral variants and some of these mutations resulted in structural changes in the S protein, thereby providing higher selection potential for increased transmission and pathogenicity of the virus.[21, 22] In BNT/BNT vaccinees, our data revealed a 5.1-fold reduction in the effectiveness of neutralizing antibodies against the B.1.617.2 VOC, thereby providing further evidence for mRNA vaccination efficacy against this variant. For B.1.351, a significant 11.5-fold reduction in neutralizing capacity was observed in vaccinees. These results agree with recent studies.[23-25]

Data of individuals that were vaccinated with ChAdOx1-S and subsequently with BNT162b2 showed a lesser reduction of nAbs targeting VOCs. This leads to the conclusion that the combination of both vaccines results in more robust immune response regarding VOC infections. Of note, the group size for the heterologous vaccination is rather small compared with the homologous mRNA vaccinated one. Therefore, the data need to be re-confirmed with a larger cohort.

The booster immunization led to a significantly stronger production of SARS-CoV-2 specific nAbs compared to the threshold after second vaccination. Of note, the median increase was even more pronounced for VOC variants. Eight months after second vaccination nAb titers were reduced by a 10-fold factor. In some individuals, for B.1.617.2 and B.1.351 variants antibody titers were even below the limit of detection of the FRNT assay.

The decline in the neutralizing serological effect in recovered patients is however more pronounced. Here, we observed a 21.5-to 34.7-fold decrease in nAb titers for patients with mild and a 34-to 75.3-fold reduction for patients with severe COVID-19 courses. In other studies, a 3.5-fold reduction against B.1.351 was reported in convalescent patients compared to the wildtype virus.[26] One third of the individuals with mild COVID-19 did not show any detectable nAbs against B.1.1.7 and B.1.351 at all.

### BNT162b2 vaccination and mucosal immune response

Generation of an intermittent protective mucosal immunity is generally accepted in COVID-19 patients undergoing natural infection and constitutes a relevant component in the suppression of pandemic SARS-CoV-2 dissemination. However, using available COVID-19 mRNA and vector-based vaccines, an immune response of mucosa associated lymphoid tissue (MALT) is highly questionable. The protection of most systemic vaccinations against mucosal infection solely relies on few circulating IgA and IgG antibodies which transudate from sera into the mucosa.[27] These results suggest that SARS-CoV-2 mRNA vaccination is not able to trigger a detectable protective mucosal immune response. However, other groups recently detected S1 IgG antibodies in saliva of vaccinated healthcare workers, which might however be attributable to undiscovered natural SARS-CoV-2 infections that occurred previously.[28] Additional development of mucosal vaccines could be crucial to improve the suppression of the pandemic spread of future potential SARS-CoV-2 VOCs.

## Conclusions

BNT162b2 mRNA vaccination provided sustainable formation of SARS-CoV-2 neutralizing antibodies in our studied cohort. To preserve detectable nAb titers after 6 months, a booster vaccination should be considered, especially for the protection against variants of concern. A heterologous vaccine regime involving ChAdOx1-S vector-based prime and BNT162b2 mRNA vaccine boost even exceeded these titers of neutralizing antibodies and might thus feature beneficial synergy. None of the studied vaccines induced detectable mucosal immune response.

## Data Availability

All relevant data are within the manuscript and its Supporting Information files.

## Author contributions and conflict-of-interest disclosure

O.N., J.W. and S.B. designed the study, performed assays, analyzed results and wrote the manuscript. A.R., J.F. and S.U. performed assays, conducted analyses and wrote the manuscript. S.L., S.K., N.K., M.B. and C.L. recruited probands/patients, critically discussed hypotheses and revised the manuscript. None of the authors declare competing conflicts of interest.

## Funding and acknowledgements

This publication was supported by the Saxon State Ministry for Science, Culture and Tourism (grant SaxoCOV) and the European Virus Archive GLOBAL (EVA-GLOBAL) project that has received funding from the European Union’s Horizon 2020 research and innovation program under grant agreement No 871029. Furthermore, the ImmunoDeficiencyCenter Leipzig received founding from the Jeffrey Modell Foundation for Primary immunodeficiency diseases.

The authors are indebted to the voluntary help of probands and patients to participate in this study. Furthermore, we would like to particularly emphasize the efforts made by Ulrike Schmidt and Cathrin Crimmann, and all participating doctors and nurses during the vaccination campaign. Moreover, we like to thank Ulrike Ehlert and Steffen Jakob for their excellent technical assistance. We also thank Corinna Pietsch (Leipzig University Hospital) and Klaus Überla (Erlangen-Nürnberg University) for supplying VOCs.

